# Development and Validation of a Machine Learning Model Integrated with the Clinical Workflow for Inpatient Discharge Date Prediction

**DOI:** 10.1101/2024.06.24.24309419

**Authors:** Mohammed A. Mahyoub, Kacie Doughetry, Ravi Yadav, Raul Berio-Dorta, Ajit Shukla

## Abstract

**Background:** Discharge date prediction plays a crucial role in healthcare management, enabling efficient resource allocation and patient care planning. Accurate estimation of the discharge date can optimize hospital operations and facilitate better patient outcomes.

**Materials and Methods:** In this study, we employed a systematic approach to develop a discharge date prediction model. We collaborated closely with clinical experts to identify relevant data elements that contribute to the prediction accuracy. Feature engineering was used to extract predictive features from both structured and unstructured data sources. XGBoost, a powerful machine learning algorithm, was employed for the prediction task. Furthermore, the developed model was seamlessly integrated into a widely used Electronic Medical Record (EMR) system, ensuring practical usability.

**Results:** The model achieved a performance surpassing baseline estimates by up to 35.68% in the F1-score. Post-deployment, the model demonstrated operational value by aligning with MS GMLOS and contributing to an 18.96% reduction in excess hospital days.

**Conclusions:** Our findings highlight the effectiveness and potential value of the developed discharge date prediction model in clinical practice. By improving the accuracy of discharge date estimations, the model has the potential to enhance healthcare resource management and patient care planning. Additional research endeavors should prioritize the evaluation of the model’s long-term applicability across diverse scenarios and the comprehensive analysis of its influence on patient outcomes.

## 1. Introduction

A key aspect of optimal healthcare delivery is the ability of hospitals and clinicians to achieve both patient-centered care and efficient resource utilization [1], [2]. These objectives are interrelated: Early hospital discharge can have positive clinical implications for patients. Evidence has shown that patients who experience prolonged hospital stays have increased susceptibility to hospital-acquired infections, pressure ulcers, and nutritional deterioration [3], [4], [5]. A potential benefit of discharging patients when they no longer require hospital-level care is the prevention of complications associated with prolonged hospitalization. Additionally, timely discharge is crucial for ensuring the availability of patient beds for those in the Emergency Department (ED) who need them [6]. The timely transfer of critically ill patients from the ED to an appropriate inpatient unit is essential for optimal patient outcomes. Previous studies have shown that prolonged ED boarding times can lead to increased hospital length of stay and higher mortality rates for these patients [7].

A phenomenon that occurs infrequently but has serious implications for patient safety is the departure of patients who need hospital care from emergency departments without being seen by a physician. This can happen when EDs are overcrowded due to inefficient patient flow processes. Patients who leave without being seen may experience adverse outcomes that could have been prevented or mitigated by timely medical attention [8].

One of the factors that affects the quality and cost of care is patient flow, which refers to the efficient use of resources and time during the patient’s stay in the hospital [9], [10]. A common practice for planning the discharge of patients is to rely on the clinicians’ estimates of the discharge date. However, this practice has some drawbacks, such as consuming the clinicians’ time that could be spent on other tasks or direct patient care, and having low accuracy [11], [12]. A possible alternative is to apply machine learning models to predict the discharge date of patients based on their length of stay (LOS) in the hospital. Several studies have proposed and developed different LOS models for this purpose [11], [13], [14], [15], [16], [17], [18]. However, most of these studies have focused on the model development stage, using retrospective data, creating features for LOS prediction, training, and evaluating different models, without considering how to implement them in practice. The majority of published articles in healthcare machine learning have failed to showcase successful implementations of proposed models, resulting in a substantial gap between theoretical concepts and practical applications. This underscores the pressing need for practical research that addresses the entire life cycle of predictive models, spanning from their initial conceptualization to their effective integration into operational systems [19], [20].

This article introduces a practical machine learning model that combines various clinical data sources, such as demographics, complaints, and medical problems, to predict the discharge date for patients. By enabling early and proactive discharge planning, this model offers significant benefits. The key contributions of this paper are as follows:

- It develops a machine learning model that is tailored for discharge date prediction and can be seamlessly integrated into the clinical workflow.
- It integrates the predictive model with a widely adopted electronic medical record system in the United States.
- It goes beyond previous studies by evaluating the model after its deployment, a step that is often overlooked in similar research, providing valuable insights into the model’s real-world performance and effectiveness.

This paper is organized as follows. Section 2 delves into the materials and methods employed in this research. It encompasses an explanation of the data sources utilized, the methods employed for data collection, and an outline of the model development and deployment process. In Section 3, the results and findings of the study are presented. This includes an analysis of the model’s performance during both the development and deployment stages, providing insights into its effectiveness and reliability. Lastly, in Section 4, the paper concludes by highlighting the main contributions and outcomes of the research, summarizing the key findings and implications derived from the study.

## 2. Materials and Methods

### 2.1. Methodology Overview

We present a systematic and rigorous approach to develop a predictive model for inpatient discharge date, as illustrated in Figure 1. Our approach consists of five phases. In the first phase, we define the healthcare problem and analyze the underlying process. In the second phase, we identify the relevant data elements and validate them with clinical teams. Then, we map the data elements to the clinical database and retrieve them for further analysis. Next, we preprocess and transform the data for modeling purposes. In the third phase, we develop and evaluate various machine learning models and select the best one to proceed to the fourth phase, where we deploy and integrate the model within the electronic medical record (EMR) system. Finally, in the fifth phase, we monitor the model’s performance after deployment, assess its operational impact and evaluate its effect on the healthcare process under investigation.

**Figure 1.**
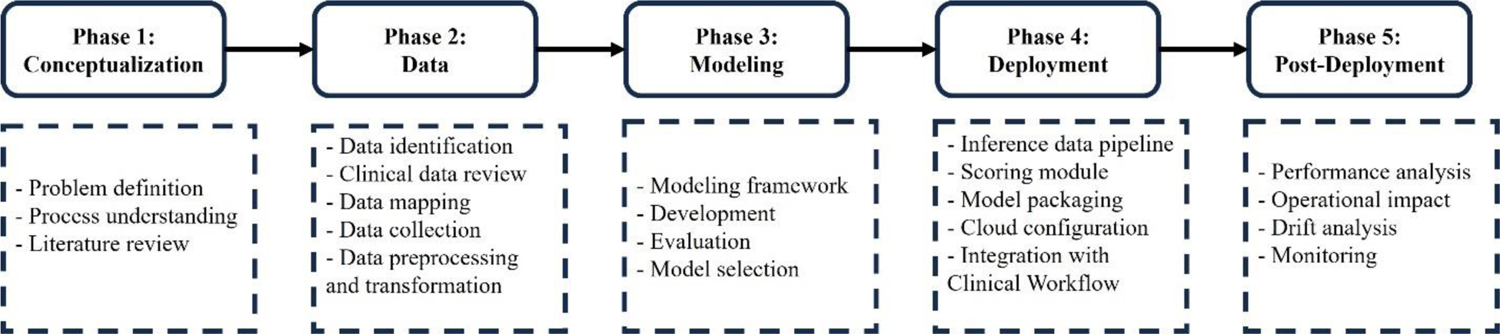
Research methodology overview.

### 2.2. Data

We identified the data based on the clinical team’s input and the relevant literature review. We then mapped the collected data to the corresponding fields and tables in our EMR system (Epic). Next, we developed an SQL query to extract the dataset from the Clarity database. We also performed a temporal analysis to select only the data elements that were available before the model run (18 hours after inpatient admission). The final list of data elements included: age, marital status, sex, admission date, admission source, problems, complaints, and MS Geometric Mean Length of Stay (MS GMLOS).

The raw data underwent a comprehensive preprocessing pipeline to prepare it for machine learning modeling. Considering the negligible proportion of missing values, rows containing any nulls were removed entirely. The categorical feature “marital status” was transformed into a binary feature, assigning a value of 1 to married patients and 0 otherwise. Sex was similarly encoded using two binary features: “Is_Male” and “Is_Female”. The admission date was utilized to create situational binary features indicating admission on Friday, Saturday, and Sunday (“Is_Friday”, “Is_Saturday”, “Is_Sunday”).

One-hot encoding was applied to the “admission source” variable, generating 27 binary features corresponding to the 27 unique admission sources. “Problems” and “complaints” underwent similar preprocessing. For each patient, all complaints were concatenated into a single text string, which was then cleaned by removing extra spaces and punctuation, converting to lowercase, and tokenizing into words. Subsequently, unique words were extracted, with each word represented by a binary feature: 1 if present in the complaint, 0 otherwise. After eliminating highly sparse features, 64 binary features remained for complaints. The same approach was applied to problems, resulting in 727 features. In total, the preprocessing steps yielded 814 features suitable for machine learning analysis.

This study focuses on the MS Geometric Mean Length of Stay (MS GMLOS) as the target variable, selected for its efficacy in streamlining discharge processes and reducing patient excess days. Unlike the arithmetic mean, prone to skewing by outliers, GMLOS offers a more accurate depiction of typical patient flow within a Diagnosis-Related Group (DRG) by calculating the geometric mean of all lengths of stay. This emphasis on central tendency makes GMLOS valuable for identifying patients at risk of prolonged stays and implementing targeted interventions for their expedited discharge. To adapt MS GMLOS for use in multi-class classification models, we categorized its continuous values into five discrete groups: discharged within 1, 2, 3, 4 days, and those with stays lasting 5 or more days. This transformation enables the application of classification algorithms to predict the likelihood of patients falling into these predefined discharge timeframes.

The dataset, following the preprocessing steps outlined above, comprised 99,561 samples. We allocated 69,692 samples for training the model, while the remaining 29,869 (30%) samples were reserved for testing the model’s performance.

### 2.3. Model Development

Using predictive modeling, this study proposes a system to estimate the discharge date of patients. The system computes a score for each patient based on their attributes, which reflects their predicted discharge date. The predictive system’s complexity can range from a simple linear or nonlinear function to a highly sophisticated one that requires additional algorithms for interpretation. As the complexity increases, the system becomes more capable of detecting intricate patterns and associations in the data. For estimating discharge dates, there is a complex and nonlinear association between the score and patient attributes, which requires the application of advanced machine learning algorithms. In particular, this study uses the Extreme Gradient Boosting algorithm (XGBoost) [21].

XGBoost stands as a contemporary algorithm in predictive modeling, leveraging the tree-boosting framework. This technique combines numerous weak learners into a robust learner, predominantly employing decision trees with constrained depth or leaf count. Sequential training of these trees aims to rectify the errors of preceding iterations, culminating in a final prediction derived through a weighted aggregation of individual tree outputs. Our study implements the XGBoost package in Python, renowned for its scalability and efficiency in tree boosting. XGBoost exhibits notable advantages over alternative boosting methodologies, including regularization, parallelization capabilities, and proficiency in handling missing data.

### 2.4. Model Deployment

To optimize the usability of the model for clinicians, we successfully integrated it into the clinical workflow. Our integration efforts involved hosting the model on the Epic Nebula, which is an Epic cloud computing platform (Epic Systems, Verona, WI, United States). Figure 2 provides an overview of this integration. To facilitate the integration process, we utilized the Epic Slate Environment to configure and thoroughly test the model artifacts. This setup ensured a smooth and reliable transfer of data between the Chronicles database and the cloud-based model for EDD generation. The resulting scores were seamlessly delivered back to the system, enabling the placement of the EDD in the patient list to be used during clinical discussions and patient flow planning. To provide input data for the model, we customized Epic workbench reports specifically 8 tailored to the discharge date prediction model. These reports served as the means through which data was supplied to the model. Additionally, we established a batch job that executed the model on an hourly basis. Through our comprehensive integration process, we successfully embedded the model within the clinical workflow, allowing for efficient data exchange, EDD scoring, and EDD integration with patient lists. This seamless integration enhances the model’s accessibility and usability for clinicians, ultimately improving patient care and outcomes.

**Figure 2.**
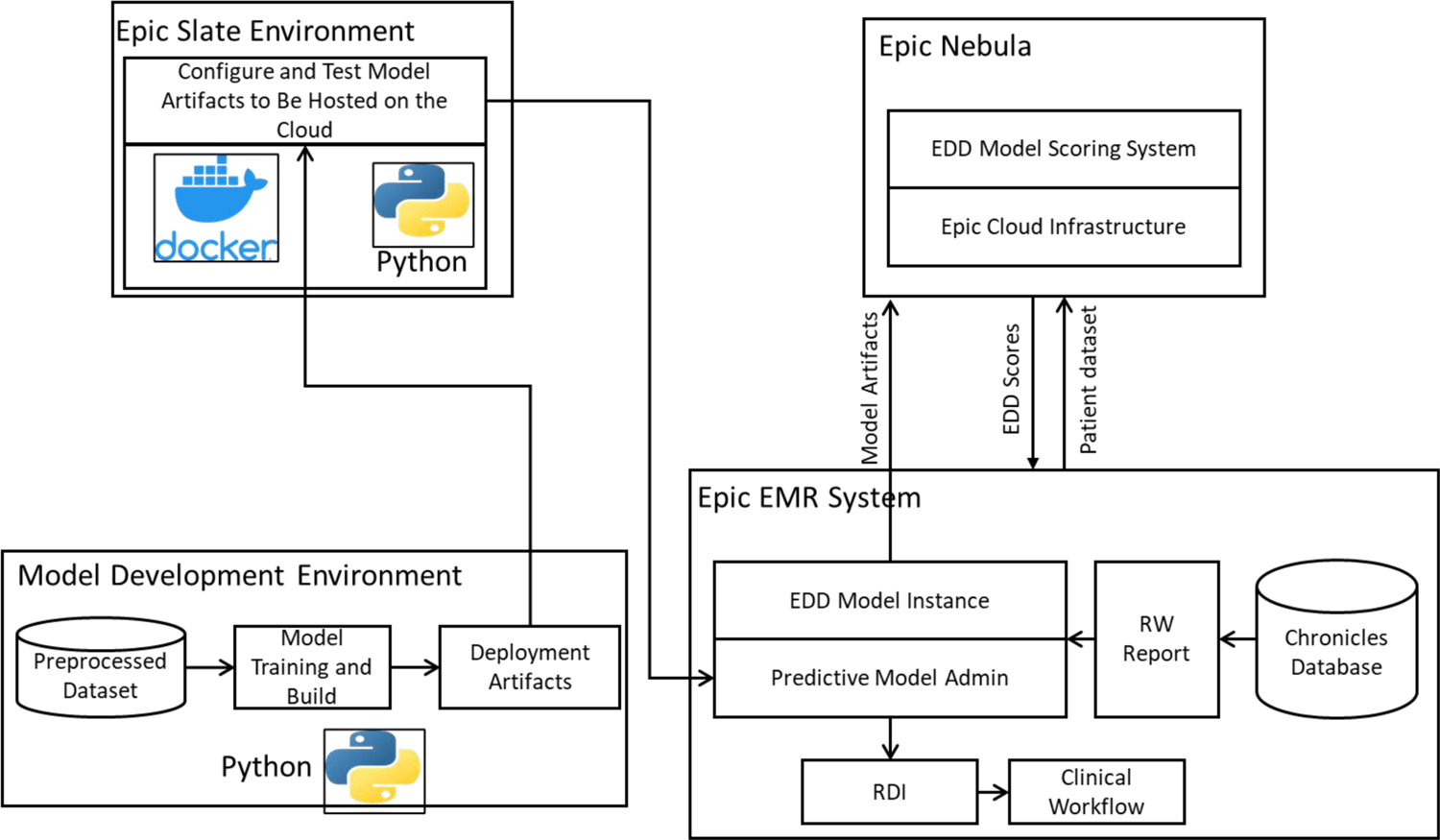
Model deployment and operationalization, adopted from [22].

### 2.5. Integration with Clinical Workflow

The model scores, specifically the estimated discharge date (EDD), were transferred from the Epic Nebula cloud back to the real-time Chronicles database. These scores were recorded in flowsheet rows and seamlessly integrated into the interdisciplinary round (IDR) lists as a distinct column labeled “Model EDD,” positioned alongside the clinical EDD column populated by clinicians. The Model EDD was generated once the data threshold was met, defined as the patient being an inpatient for 18 hours or more. The Model EDD is generated once per patient when their stay exceeds 18 hours. However, the current model configuration does not update the EDD thereafter.

## 3. Results and Discussion

In this section, we will delineate the results and findings of our research. Initially, we will evaluate the model following its development. Subsequently, we will present the results of post-deployment performance. Lastly, we will highlight the operational outcomes that have emerged from the model’s practical implementation.

### 3.1. Model Evaluation

In this study, we tackled a multi-class classification problem (five classes) to predict the estimated discharge date. The model was evaluated on the hold-out testing dataset comprising 29,869 samples. We assessed the model’s performance using the F1-score metric (see Table 1). All evaluations were compared against two baseline predictions:

- Predicting a discharge date of three days (patient is discharged in 3 days from admission) for each patient, which represents the legacy process for estimating discharge dates before deploying the predictive model.
- Randomly assigning a class to a patient, where each class (representing the estimated discharge date) is assigned with an equal likelihood of one-fifth.

**Table 1.** Evaluation metrics.

| Metric | Formula |
| --- | --- |
| Recall | $\text{Precision}_i = \frac{T_i}{T_i + F_i}$<br>where $T_i$ is the number of correctly predicted samples for class $i$ (true positives), and $F_i$ is the sum of incorrectly predicted samples that were predicted as class $i$ (false positives). |
| Precision | $\text{Recall}_i = \frac{T_i}{T_i + M_i}$<br>where $T_i$ is the number of correctly predicted samples for class $i$ (true positives), and $M_i$ is the sum of actual samples of class $i$ that were incorrectly predicted as other classes (false negatives). |
| F1-Score | $\text{F1-Score}_i = 2 \times \frac{\text{Precision}_i \times \text{Recall}_i}{\text{Precision}_i + \text{Recall}_i}$ |

The F1-score is a widely recognized metric for evaluating classification models, representing the harmonic mean of precision and recall. Precision, also referred to as the positive predictive value, denotes the proportion of true positives among all positive predictions. Recall, or sensitivity, assesses the model’s ability to accurately identify positive classes. Together, these metrics offer a comprehensive evaluation of the model’s predictive performance. Figure 3 compares the baseline F1 scores to those of the predictive model.

**Figure 3.**
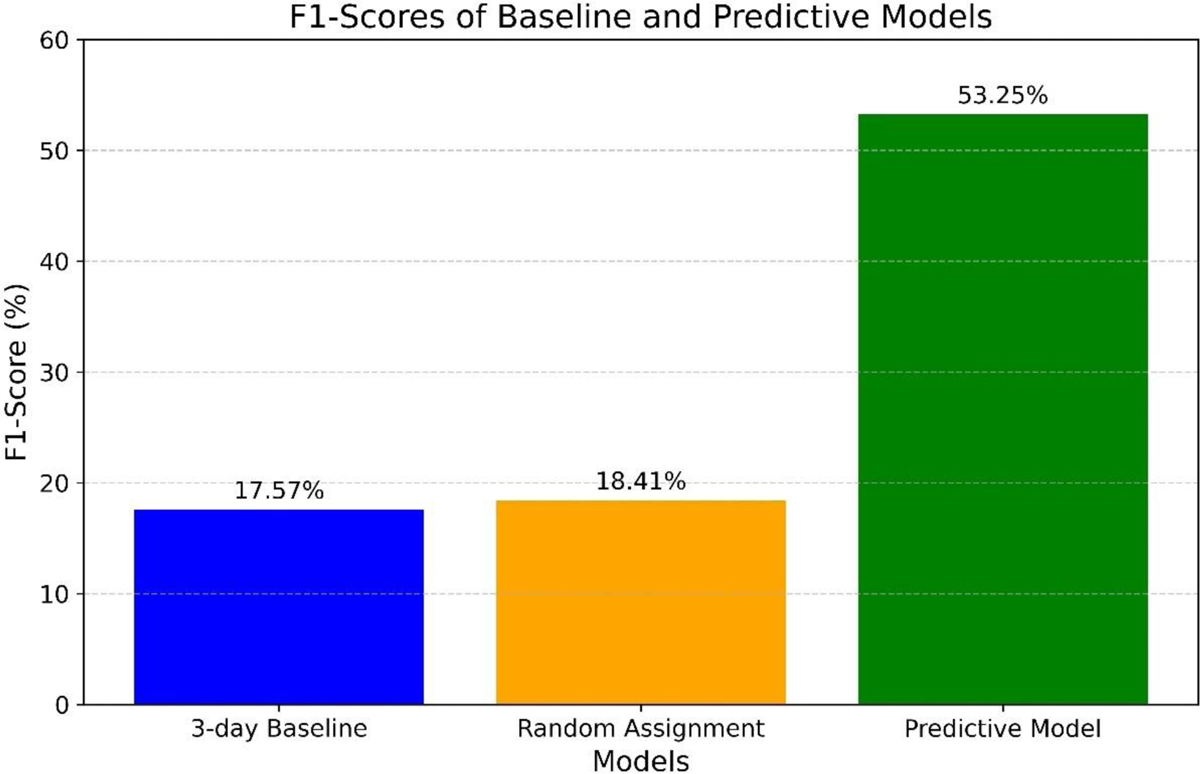
Model pre-deployment evaluation: F1-Score.

The 3-day baseline F1-score is 17.57%, signifying a very low score with minimal utility for solving the classification problem. The random label assignment method achieved an F1-score of 18.41%. In contrast, the predictive model attained an F1-score of 53.25%, demonstrating significant improvement compared to the baseline scores, particularly with a 35.68% gain over the current estimation process (estimating three days for all patients). This improvement can prove instrumental in optimizing patient flow and reducing excess days, as will be discussed later.

Several factors may have influenced the observed model F1 score. First, the dataset was limited to a few demographic details, problem lists, admission sources, and complaints. Expanding the dataset to include additional variables like vital signs, laboratory data, and medications could improve model performance. Second, the data collection window is restricted to 18 hours post-admission to facilitate early prediction, which may exclude nuanced clinical data emerging later during the patient’s hospital journey. It is argued that dynamically incorporating more clinical data elements and generating model scores more frequently during the patient’s stay would likely improve evaluation scores as the patient’s discharge time approaches. Moreover, predicting the discharge date is a multifaceted issue that extends beyond clinical factors alone. Workflow processes, clinical team decisions, transportation, and other considerations also influence discharge date determination. Collecting data on these aspects can be challenging. Hence, further investigations are recommended to explore the issue from these perspectives.

### 3.2. Post-deployment performance

In this study, we assess the post-deployment performance of the model with respect to addressing the original goal of reducing excess days relative to the MS Geometric Mean Length of Stay (MS GMLOS).

We revisit the F1 score to gauge the model’s operational performance and measure any drift from its pre-deployment performance. As illustrated in Figure 4-a, the model’s overall F1 score is 44.24% (based on operational data collected from January 2023 to April 2024, totaling 52,885 samples). The model still demonstrates advantages over the previously used approach of assigning a standardized three-day estimated inpatient length of stay. However, a significant 9% drop in the F1 score was observed compared to pre-deployment. The F1 score remained relatively stable at around 44% (see Figure 4-b).

**Figure 4.**
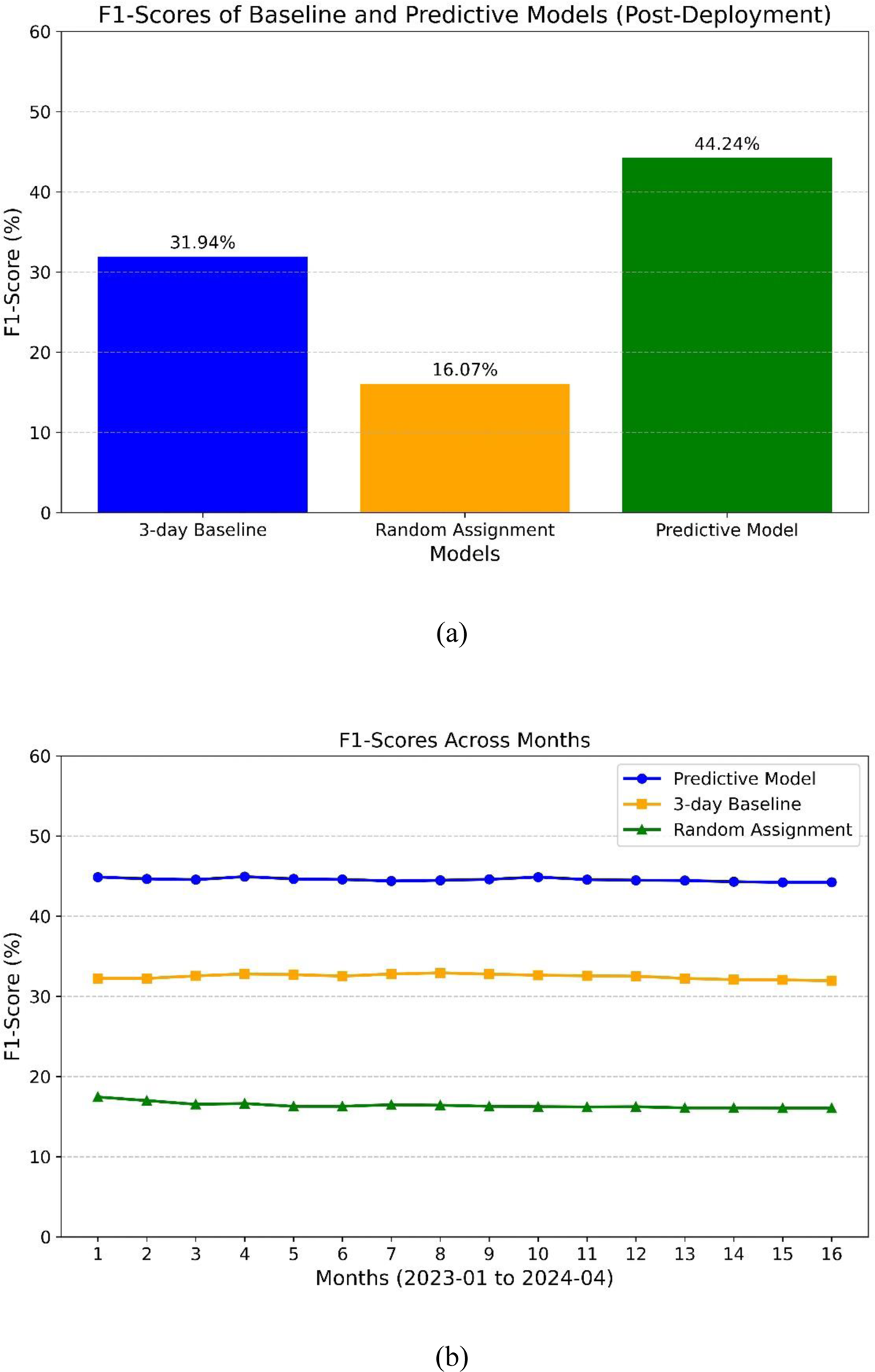
Post-deployment F1-scores.

The decrease in performance can be attributed to several factors. Firstly, the operational inference data pipeline pulls data from a real-time database, while the training data during development was collected from a backend offline database. Although efforts were made to align the data between the two environments, some unaccounted-for variables may still warrant further investigation. Besides potential data drift, other external factors, such as modifications to clinical workflows in response to the model’s deployment, might have influenced the results. Nevertheless, the model’s utility in operations remains evident, as will be further discussed.

A crucial aspect of evaluating the model’s operational impact is to assess how well it aligns with the target benchmark (MS GMLOS) to minimize excess days. In Figure 5, we compare the average LOS for the predictive model, MS GMLOS, initial clinical estimation-based LOS, and actual LOS. Data was clipped to a range of 1 to 5 days to accommodate the fact that the predictive model is a multi-class classification model, providing five possible discharge durations: one day, two days, and so on.

**Figure 5.**
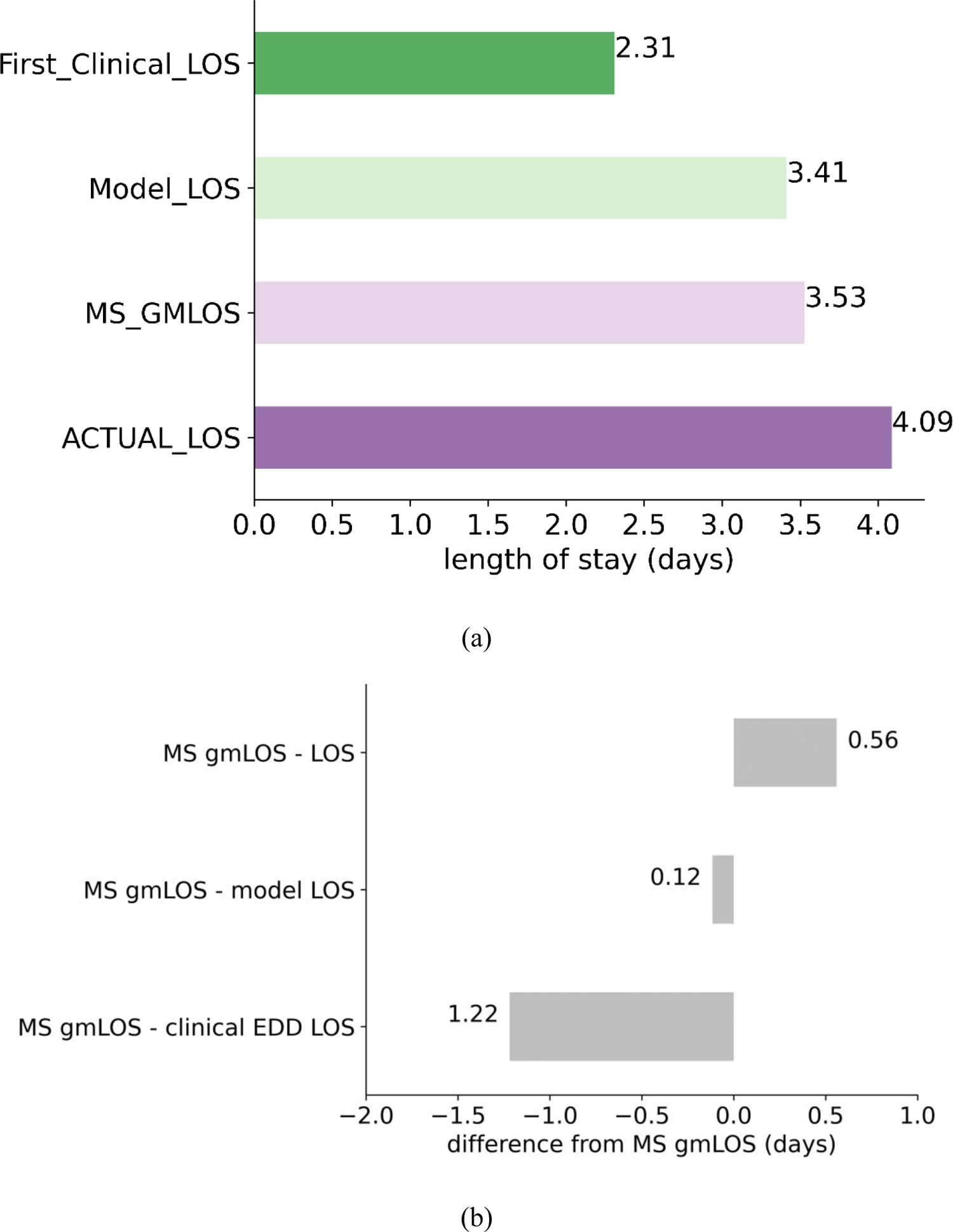
The average length of stay comparison.

The predictive model aligns closely with the MS GMLOS, with a slight underestimation (difference of 0.12 days). In contrast, the initial clinical estimation-based LOS significantly underestimates, with a 1.22-day shortfall compared to MS GMLOS. Such underestimation can disrupt the patient discharge planning process. Conversely, the actual LOS overestimates by 0.56 days, leading to excess days. The model’s alignment with the MS GMLOS can potentially help reduce excess days and optimize the patient discharge planning process.

The operational utility of the model can also be assessed based on the number of predictions that are less than or equal to the MS GMLOS, exceed it by one day, or exceed it by two or more days (refer to Table 2). In 75.14% of cases, the model predicted an LOS that was less than or equal to the GMLOS, prompting the clinical team to initiate proactive discharge planning and potentially reducing excess days. This highlights the model’s effectiveness in aligning with the desired discharge benchmark. By closely predicting the GMLOS, the model aids healthcare providers in managing patient flow more efficiently and helps in timely discharge planning. The model’s ability to predict LOS within the benchmark range supports the goal of reducing excess hospital days and optimizing resource utilization, which is crucial in improving hospital efficiency and patient outcomes.

**Table 2.** Proportions relative to MS GMLOS.

| **Criteria** | **Proportion (%)** |
| --- | --- |
| Less than or equal to MS GMLOS | 75.14 |
| Larger than MS GMLOS by 1 day | 11.73 |
| Larger than MS GMLOS by 2 days or more | 13.13 |

### 3.3. Operational Outcomes

In this analysis, we compare the excess days recorded in 2022 with those in 2023, during which the model was actively in operation. As illustrated in Figure 6, there was a substantial reduction of 13,883 days (18.96%) in excess days between 2022 and 2023. The predictive model played a significant role, along with other accompanying technological implementations, in reducing these excess days.

**Figure 6.**
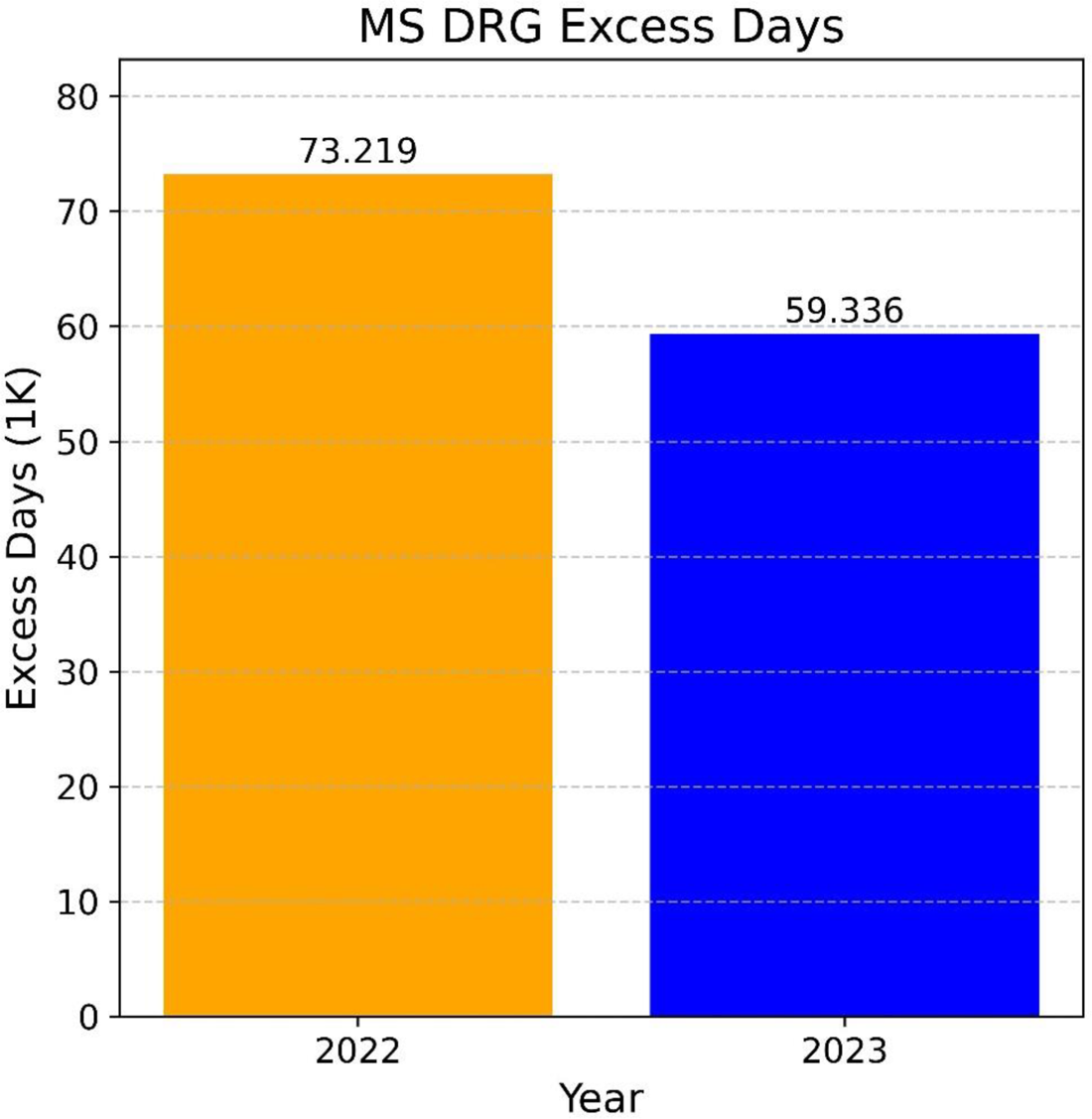
MS DRG excess days.

This reduction underscores the positive impact of data-driven decision-support tools on hospital management. Such tools help streamline discharge planning and enhance resource utilization by providing healthcare professionals with data-driven insights. By accurately forecasting patient length of stay, these models enable better scheduling and utilization of hospital resources, resulting in improved patient care and increased hospital efficiency. The reduction in excess days not only minimizes unnecessary costs associated with prolonged hospital stays but also enables hospitals to accommodate more patients, effectively improving the overall healthcare delivery system. The predictive model’s ability to integrate seamlessly into the clinical workflow demonstrates the transformative potential of advanced technology in addressing critical challenges in healthcare operations.

### 3.4. Operational and Clinical Implications

Accurate estimation of patient discharge date facilitates ancillary services, such as Social Work and Outcomes Management, to initiate the process of insurance authorization for necessary post-discharge services like subacute rehabilitation. This proactive approach prevents delays in patient discharge due to pending service authorizations and contributes to a reduction in excess hospital days.

For families, the predictive model enhances the accuracy of discharge estimations, thereby improving communication and preparation for the patient’s transition from the hospital. Knowing the estimated discharge date in advance allows families to prepare adequately for the patient’s return home or transfer to another facility, such as subacute rehabilitation. This foresight enables Outcomes Management, nurses, and physicians to engage in discussions about patient disposition with the family several days before discharge, rather than on the day of discharge. Consequently, these early conversations foster better and more effective communication between the care team and the patient’s family, ensuring a smoother transition and reducing stress for all parties involved.

## 4. Conclusions

This study illustrates the development and deployment of a machine-learning model for inpatient discharge date prediction. By leveraging XGBoost and integrating the model into the clinical workflow through Epic EMR, the research offers a practical and scalable approach to enhancing patient flow and resource management in hospitals. The results demonstrate significant improvements in reductions of excess days, highlighting the operational benefits of the model. Despite challenges like data drift and limitations in data scope, the study shows how machine learning models can provide actionable insights to improve hospital efficiency and patient outcomes. Further research is needed to refine the model by incorporating more clinical data and dynamic features, and to evaluate its long-term applicability across different healthcare settings.

## Ethics statement

The studies involving humans were approved by Virtua Health Institutional Review Board FWA00002656. The studies were conducted in accordance with the local legislation and institutional requirements. The ethics committee/institutional review board waived the requirement of written informed consent for participation from the participants or the participants’ legal guardians/next of kin because the research involved no more than minimal risk to subjects, could not practically be carried out without the waiver, and the waiver will not adversely affect the rights and welfare of the subjects. This requirement of consent was waived on the condition that, when appropriate, the subjects will be provided with additional pertinent information about participation.

## Data Availability

The data analyzed in this study is subject to the following licenses/restrictions: data in the present study are not available due to agreements made with the IRB of Virtua Health. Requests to access these datasets should be directed to Ajit Shukla.

## Notes

### Competing Interest Statement

The authors have declared no competing interest.

### Funding Statement

This study did not receive any funding.

### Author Declarations

Virtua Health Institutional Review Board (FWA00002656) gave ethical approval for this work. Protocol ID: G23109

